# Immunogenicity of BNT162b2 Vaccine Booster Dose in Patients with Inflammatory Bowel Disease Receiving Infliximab Combination Therapy: A Prospective Observational Study

**DOI:** 10.1101/2022.05.03.22274608

**Authors:** Mohammad Shehab, Fatema Alrashed, Ahmad Alfadhli, Abdulwahab Alsayegh, Usama Aldallal, Preethi Cherian, Irina Alkhair, Thangavel Alphonse Thanaraj, Arshad Channanath, Ali A. Dashti, Anwar Albanaw, Hamad Ali, Mohamed Abu-Farha, Jehad Abubaker, Fahd Al-Mulla

**Affiliations:** Division of Gastroenterology, Department of Internal Medicine, Mubarak Alkabeer University Hospital, Kuwait University, Kuwait; Department of Pharmacy Practice, Faculty of Pharmacy, Health Sciences Center (HSC), Kuwait University, Jabriya, Kuwait; School of Medicine, Royal College of Surgeons in Ireland, Medical University of Bahrain, Kingdom of Bahrain; Department of Biochemistry and Molecular Biology, Dasman Diabetes Institute (DDI), Dasman, Kuwait; Department of Genetics and Bioinformatics, Dasman Diabetes Institute (DDI), Dasman, Kuwait; Department of Medical Laboratory Sciences, Faculty of Allied Health Sciences, Health Sciences Center (HSC), Kuwait University, Jabriya, Kuwait

**Keywords:** IBD, Vaccine, Infliximab, COVID-19, immunogenicity

## Abstract

**Introduction:** Few data exist regarding the immunogenicity of third dose of BNT162b2 relative to second dose in patients with inflammatory bowel disease (IBD) on different immunosuppressive therapies. We investigated the immunogenicity of BNT162b2 vaccine booster dose in patients with IBD on infliximab combination therapy.

**Methods:** This is prospective single center observational study conducted between January 1st, 2022 until February 28th, 2022. Patients were recruited at the time of attendance at the infusion center. Eligibility criteria included patients with confirmed diagnosis of IBD who are receiving infliximab with azathioprine or 6-mercaptopurine and have received two or three-dose of BNT162b2 vaccine. Patients were excluded if they were infected or had symptoms of SARS-CoV-2 previously since the start of the pandemic or received other vaccines than the BNT162b2. Our primary outcome was the concentrations of SARS-CoV-2 antibodies Immunoglobulin G (IgG) and neutralizing antibodies 40-45 weeks from the first dose of BNT162b2 in patients with IBD receiving infliximab combination therapy. Medians with interquartile range (IQR) were calculated.

**Results:** 162 patients with IBD and receiving infliximab combination therapy were recruited and the number of patients in each group was 81. Median (IQR) SARS-CoV-2 IgG levels were significantly lower after the second dose [125 BAU/mL (43, 192)] compared to patients who received the third booster dose [207 BAU/mL (181, 234)] (p = 0.003). Neutralizing antibody levels were also lower after the second dose [80 BAU/mL (21, 95)] compared to patients who received the third booster dose [96 BAU/mL (93, 99)] (p = <0.001). The percentage of patients who achieved positive SARS-CoV-2 IgG levels in the third (booster) dose group was higher (96.3%) than those in second dose group (90%)(p = 0.026). Percentage of patients who received third (booster) dose and achieved positive SARS-CoV-2-neutralizing antibody level was 100%, whereas it was lower (88.9%) in patients who received second dose only (p=0.009).

**Conclusion:** Most patients with IBD on infliximab combination therapy had positive SARS-CoV-2 IgG and neutralizing antibody concentrations 40-45 weeks post BNT162b2 vaccination. However, SARS-CoV-2 IgG and neutralizing antibody concentrations were lower in patients who received 2 doses only compared to patients who received a third dose.

## Introduction

Coronavirus was first identified to have infected people in the mid-1960s. Since then, multiple subgroupings have emerged. Recent and most notable of which have been severe acute respiratory syndrome coronavirus 2 (SARS-CoV-2) originating from Wuhan, China.^1,2^ SARS-CoV-2 is particularly dangerous in part due to its heterogeneous symptoms resulting in anything between diarrhea to severe respiratory distress and its high infectious rate. The transmission of the virus is accomplished via the presence of angiotensin-converting enzyme 2 (ACE2) receptors, present on epithelial type II cells in the lungs and the brush border of gut enterocytes. Here, the virus can then utilize these receptors to access the host’s tissue resulting in infection.^3^ As a result of its rapid spread, SARS-CoV-2, as of January 30, 2022, has infected 364,191,494 confirmed cases, of which 5,631,457 resulted in deaths.^4^ Despite new variants like Delta and Omicron continuing to cause a global surge in cases, the advent of vaccines, including boosters, brings confidence that the spread will be curbed.

Global efforts led to development of highly effective COVID-19 vaccines, with early findings reporting 95% efficacy of mRNA vaccine BNT162b2 against COVID-19.^5,6^ Since then, however, variants sprouted, and immunity dwindled. This resulted in the reported efficacy of BNT162b2 against Omicron dropping to 34-37% after 15 weeks of the second dose. Thankfully, the arrival of boosters for BNT162b2 primary course recipients helped push COVID-19 vaccine efficacy back over 70%, making it especially important for immunocompromised patients such as those with inflammatory bowel disease (IBD).^7^

IBD is a chronic immune-mediated inflammatory process often distinguished between two subtypes, Crohn’s disease (CD) and Ulcerative Colitis (UC), impacting millions of patients. IBD is often treated using immunosuppressive drugs like corticosteroids, tumor necrosis factor inhibitors, thiopurines, and Janus kinase inhibitors raising concerns over the risk of patient complications from debilitating and/or infectious sources. This is exceptionally significant today, as recent evidence suggests that corticosteroid and 5-aminosalicylic acid (5-ASA) use is associated with more severe COVID-19 outcomes such as hospitalization and death.^8–11^ This can be explained due to a blockage of intracellular signals needed for the host to fight pathogens with patients on immunosuppressive drugs.^12,13^ Furthermore, early findings in patients receiving biological treatments such as infliximab combination therapy demonstrated lower SARS-CoV-2 IgG, IgA, and neutralizing antibody levels after BNT162b2 vaccination compared with healthy participants. All things considered, most patients displayed significant immunogenicity after two doses of the vaccines, illustrating the potential greater benefits of a third dose.^14^ Thus, this study aims to assess the immunogenicity of the second dose compared to third (booster) dose of BNT162b2 vaccine in patients with IBD receiving infliximab with azathioprine or 6-mercaptopurine (infliximab combination).

## Material and Methods

A prospective single center observational study was conducted at a tertiary care inflammatory bowel disease (IBD) center, Mubarak Al-Kabeer Hospital. Patients were recruited at the time of attendance at the infusion center between January 1st, 2022 until February 28th, 2022. This study was performed and reported in accordance with Strengthening the Reporting of Observational Studies in Epidemiology (STROBE) guidelines.^15^ The international classification of diseases (ICD-10 version:2016) was used to make diagnosis of IBD. Patients were considered to have IBD when they had ICD-10 K50, K50.1, K50.8, K50.9 corresponding to Crohn’s disease (CD) and ICD-10 K51, k51.0, k51.2, k51.3, k51.5, k51.8, k51.9 corresponding to ulcerative colitis (UC).^16^

Patients were eligible to be included if they : 1) had confirmed diagnosis of inflammatory bowel disease before the start of the study 2) were receiving infliximab with azathioprine or 6-mercaptopurine for at least 6 weeks or more for induction of remission or with at least one dose of drug received for maintenance of remission in the previous 8 weeks before first dose of vaccination 3) Second dose group: have received two-dose of COVID-19 vaccination with BNT162b2 vaccine, 3 weeks apart. Third dose (booster) group: have received two-dose of COVID-19 vaccination with BNT162b2 vaccine, 3 weeks apart and a third (booster) dose 24 weeks after the second dose (figure 1) 4) were eat least are 18 years of age or older. SARS-COV-2 PCR was performed within 72 hours of each vaccine dose and, if positive, patients were excluded. Patients were Also excluded if they were infected or had symptoms of SARS-CoV-2 previously since the start of the pandemic. In addition, patients who received other vaccines than the BNT162b2 were excluded. Patients who received corticosteroids two weeks before the first dose of the vaccine up to the time of recruitment were also excluded. Finally, patients taking other immunomodulators such as methotrexate were also excluded.

**Figure1.**
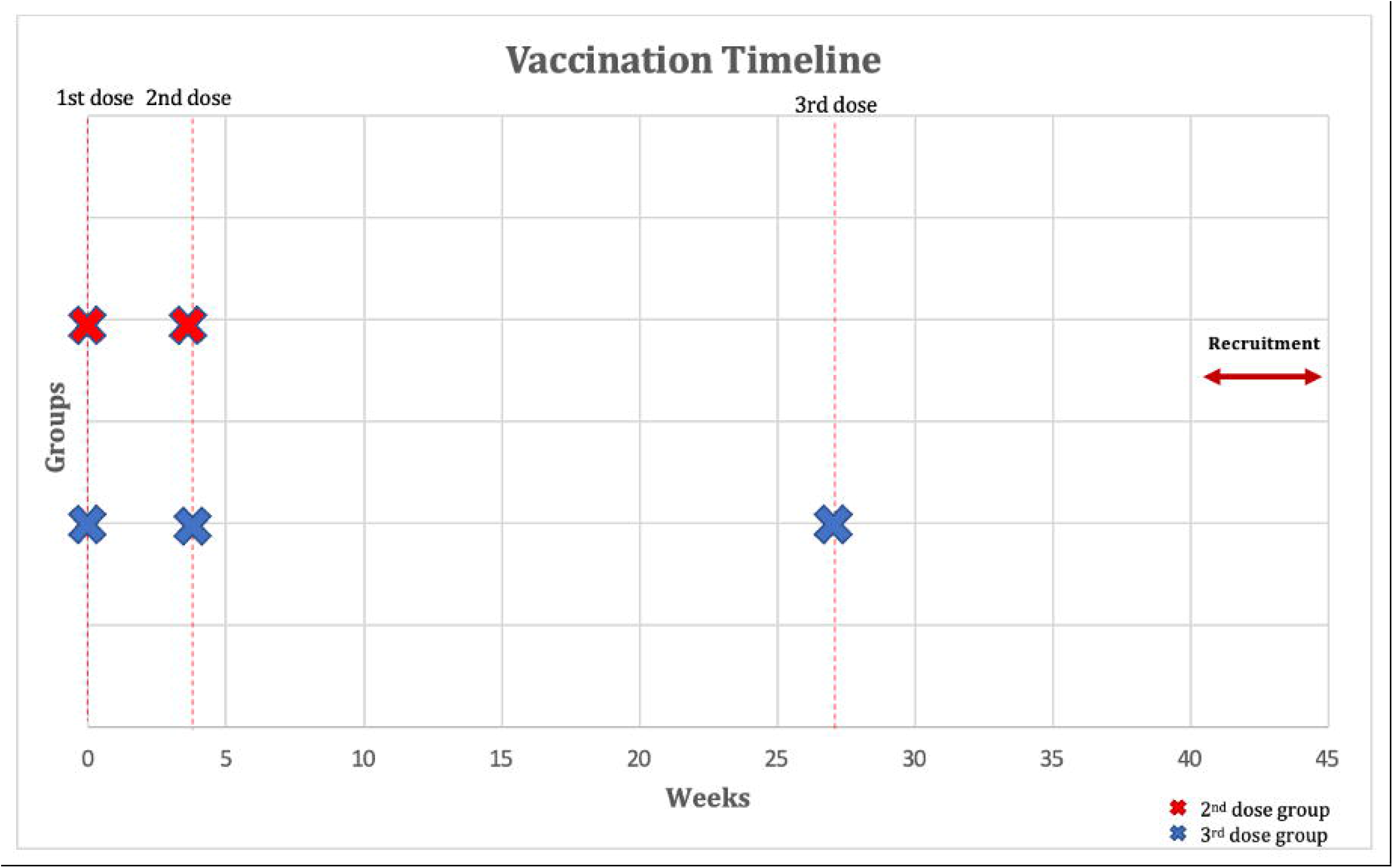
Outline of vaccination timeline and time when serological response was assessed

### Outcome measures

Our primary outcome was the concentrations of SARS-CoV-2 antibodies including Immunoglobulin G (IgG) and neutralizing antibodies 40-45 weeks after their first dose of BNT162b2 in patients with IBD receiving infliximab combination therapy. Data regarding type and extent of IBD as well as duration of infliximab combination therapy were also recorded.

### Laboratory Methods

Enzyme-linked immunosorbent assay (ELISA) kit (SERION ELISA agile SARS-CoV-2 IgG and IgA SERION Diagnostics, Würzburg, Germany) was used to measure plasma levels of SARS-CoV-2-specific IgG antibodies based on the manufacturers protocol. Units of IgG levels were reported as binding antibody units (BAU)/mL. Values of below 31.5 BAU/mL were considered negative or nonprotective. Neutralizing antibody levels below 20% were considered negative or nonprotective. The positive and negative thresholds were determined as per manufacturer’s instructions. Results were construed by calculating inhibition rates for samples as per the following equation: Inhibition = (1 - O.D. value of sample/O.D. value of negative control) ×100%.

### Ethical consideration

This study was reviewed and approved by the Ethical Review Board of Dasman institute “Protocol # RA HM-2021-008” as per the updated guidelines of the Declaration of Helsinki (64th WMA General Assembly, Fortaleza, Brazil, October 2013) and of the US Federal Policy for the Protection of Human Subjects. The study was also approved by the Ministry of Health of Kuwait (reference: 3799, protocol number 1729/2021). Subsequently, patient informed written consent was obtained before inclusion in the study.

### Statistics

We performed descriptive statistics to characterize both the second and third dose (booster) dose groups. Standard descriptive statistics were used to present the demographic characteristics of patients included in this study and their measured antibody levels. Analysis was conducted in R (R Core Team, 2017). Data are expressed as medians with interquartile range (IQR) unless otherwise indicated. Categorical variables were compared using the Fisher’s exact test or Pearson’s Chi-squared test, and continuous variables were compared with the Kruskal–Wallis rank sum test or Wilcoxon rank sum test. P-value of less than or equal to 0.05 considered statistically significant. Both Groups were matched for age, sex and time-since-first vaccine-dose using Optimal pair matching method. The technique attempts to choose matches that collectively optimize an overall criterion. The criterion used was the sum of the absolute pair distances in the matched sample. In addition, percentage of positive IgG and neutralizing antibody levels was calculated in both groups. χ2 tests were used to assess whether the percentages of positive antibodies differed across categories of both groups.

## Results

### Baseline Cohort Characteristics

Patients were recruited between 1 January 2022 and 28 February 2022. In total, 162 patients were recruited and serology assays to quantify SARS-CoV-2 antibody levels were performed for all patients. The number of patients included in both the second dose group and third (booster) dose group was 81. Mean age was 35 years old in both groups and body mass index (BMI) was lower in the second dose group compared to third dose group (25.8 vs. 24.7 kg/m^2^). In both groups, most patients had Crohn’s disease (>56%) and more than 20% pf patients were smokers. The mean duration between the serology test and last dose of vaccine was 7 (±2) weeks. The median duration of infliximab com-bination therapy was 12 months (See Table 1).

**Table 1.** Baseline characteristics of participants.

| Characteristic | Second Dose, N<br>= 81 | Booster Dose, N<br>= 81 |
| --- | --- | --- |
| Mean Age (years) | 35.2 | 35.6 |
| **Gender n (%)** |  |  |
| Female | 38 (47%) | 36 (45%) |
| Male | 43 (53%) | 45 (55%) |
| BMI (Median) | 24.7 | 25.8 |
| Smoking n (%) | 19 (23.0%) | 17 (21.0%) |
| **Co-morbidities n (%)** |  |  |
| Diabetes | 3 (3.7%) | 3 (3.6%) |
| Hypertension | 7 (8.6%) | 6 (7.1%) |
| Heart Disease | 5 (6.1%) | 6 (7.1%) |
| Arthritis or any autoimmune disease | 8 (9.8%) | 9 (11%) |
| COPD | 1 (1.2%) | 1 (1.2%) |
| Kidney Disease | 2 (2.4%) | 0 (0%) |
| Asthma | 2 (2.5%) | 1 (3.6%) |
| Hyperlipidemia | 9 (11%) | 9 (11%) |
| Duration of infliximab combination therapy (median, months) | 12 | 13 |
| **Disease Extent n (%)** |  |  |
| Ulcerative colitis (UC) | 33 (41%) | 36 (44%) |
| E1: ulcerative proctitis | 5 (16%) | 6 (18%) |
| E2: left sided colitis | 11 (32%) | 12 (33%) |
| E3: extensive colitis | 17 (52%) | 18 (49%) |
| Crohn's disease (CD) | 48 (59%) | 45 (56%) |
| L1: ileal | 26 (54%) | 23 (51%) |
| L2: colonic | 5 (10%) | 5 (11%) |
| L3: ileocolonic | 17 (36%) | 15 (34%) |
| L4: upper gastrointestinal | 0 (0%) | 2 (4%) |
| B1: inflammatory | 21 (44%) | 19 (43%) |
| B2: stricturing | 13 (28%) | 12 (27%) |
| B3: penetrating | 14 (30%) | 14 (30%) |
| **Lab Parameters** |  |  |
| CRP, mg/L (median) | 6.3 | 6.2 |
| Albumin, g/L (median) | 40 | 42 |
| ESR, mm/h | 11 | 9 |
| Stool fecal calprotectin, ug/g (median) | 114 | 112 |

### Outcomes

Median (IQR) SARS-CoV-2 IgG level was significantly lower in patients treated with infliximab combination therapy after the second dose [125 BAU/mL (43, 192)] compared to patients who received the third booster dose [207 BAU/mL (181, 234)] following vaccination with BNT162b2 (p = 0.003) (see figure 2). Neutralizing antibody levels were also lower in patients treated with infliximab combination therapy after the second dose [80 BAU/mL (21, 95)] compared to patients who received the third booster dose [96 BAU/mL (93, 99)] following vaccination with BNT162b2 (p = <0.001). (See table 2 and figure 3).

**Figure 2.**
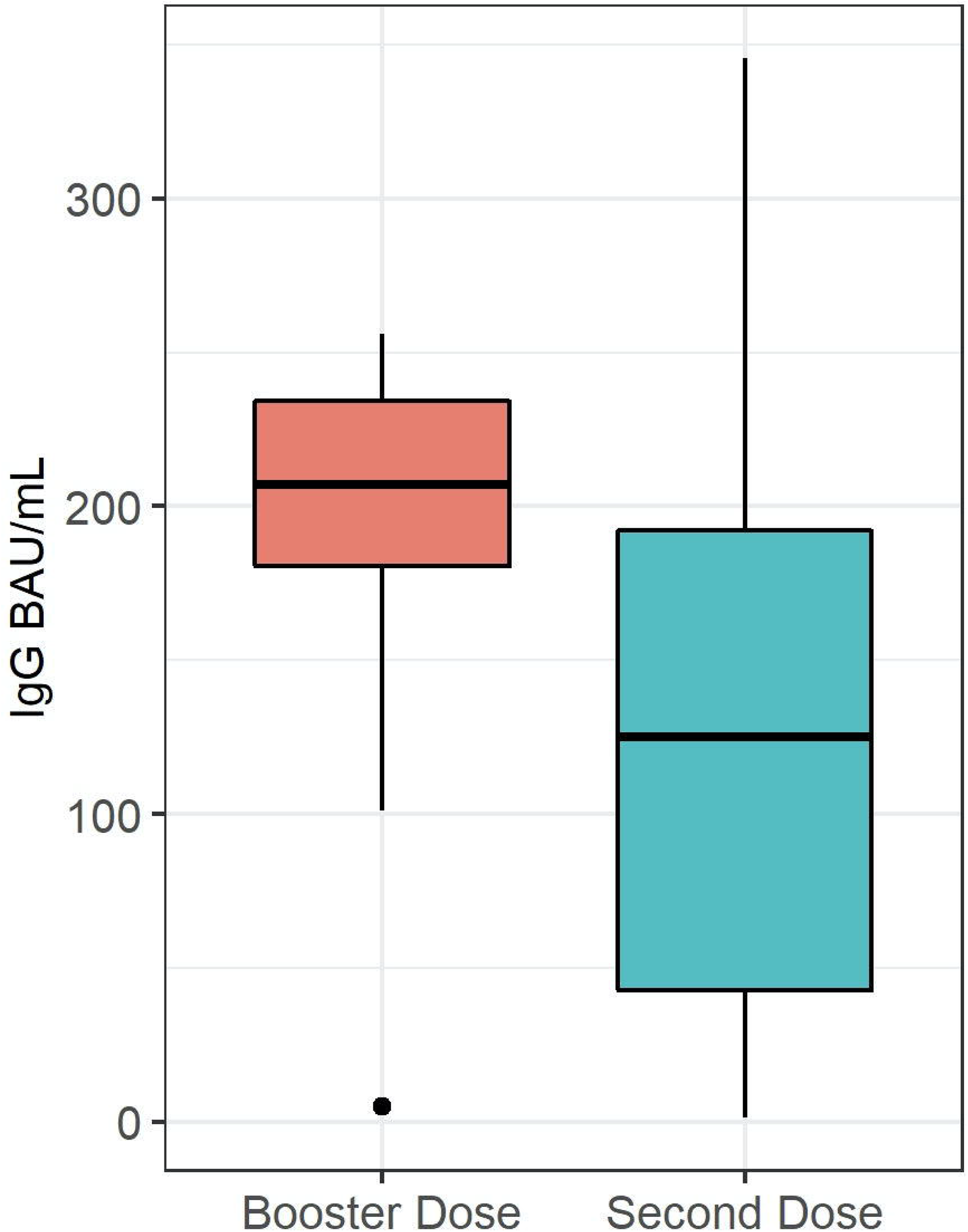
Box plot illustrating SARS-CoV-2 IgG antibody concentrations in the infliximab combination therapy after second and third dose.

**Table 2.**
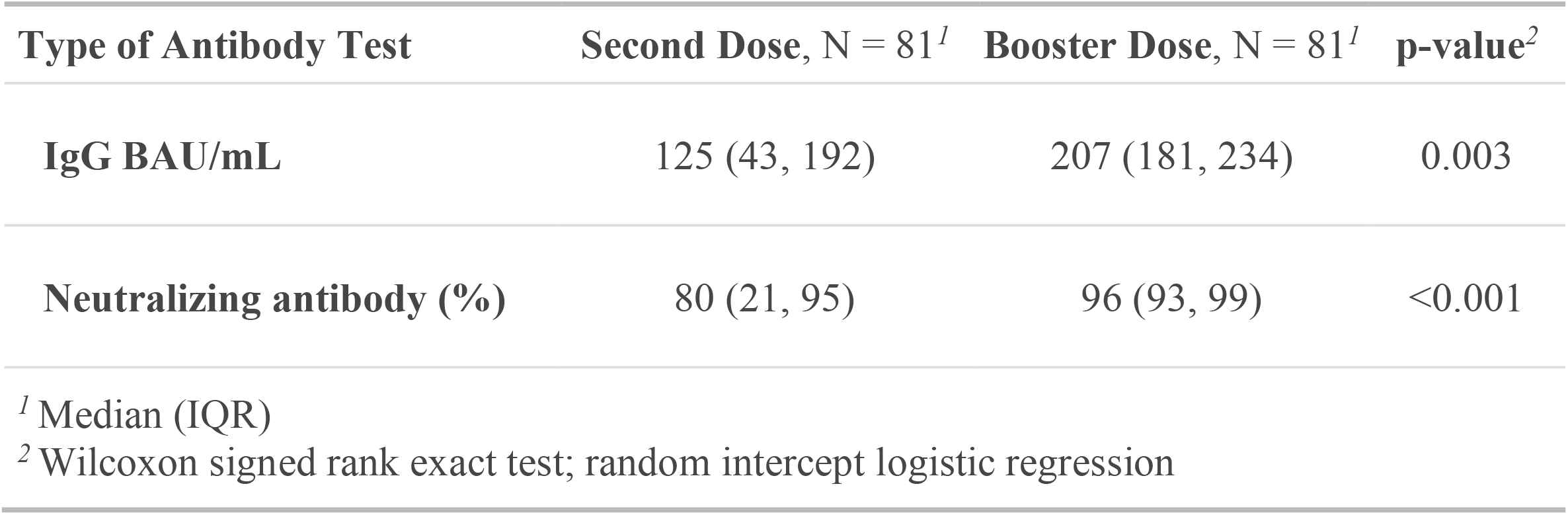
Antibody responses in patients receiving infliximab combination therapy after booster dose vs. second dose.

| Type of Antibody Test | Second Dose, N = 81<sup>1</sup> | Booster Dose, N = 81<sup>1</sup> | p-value<sup>2</sup> |
| --- | --- | --- | --- |
| **IgG BAU/mL** | 125 (43, 192) | 207 (181, 234) | 0.003 |
| **Neutralizing antibody (%)** | 80 (21, 95) | 96 (93, 99) | <0.001 |

<sup>1</sup> Median (IQR)
<sup>2</sup> Wilcoxon signed rank exact test; random intercept logistic regression

**Figure 3.**
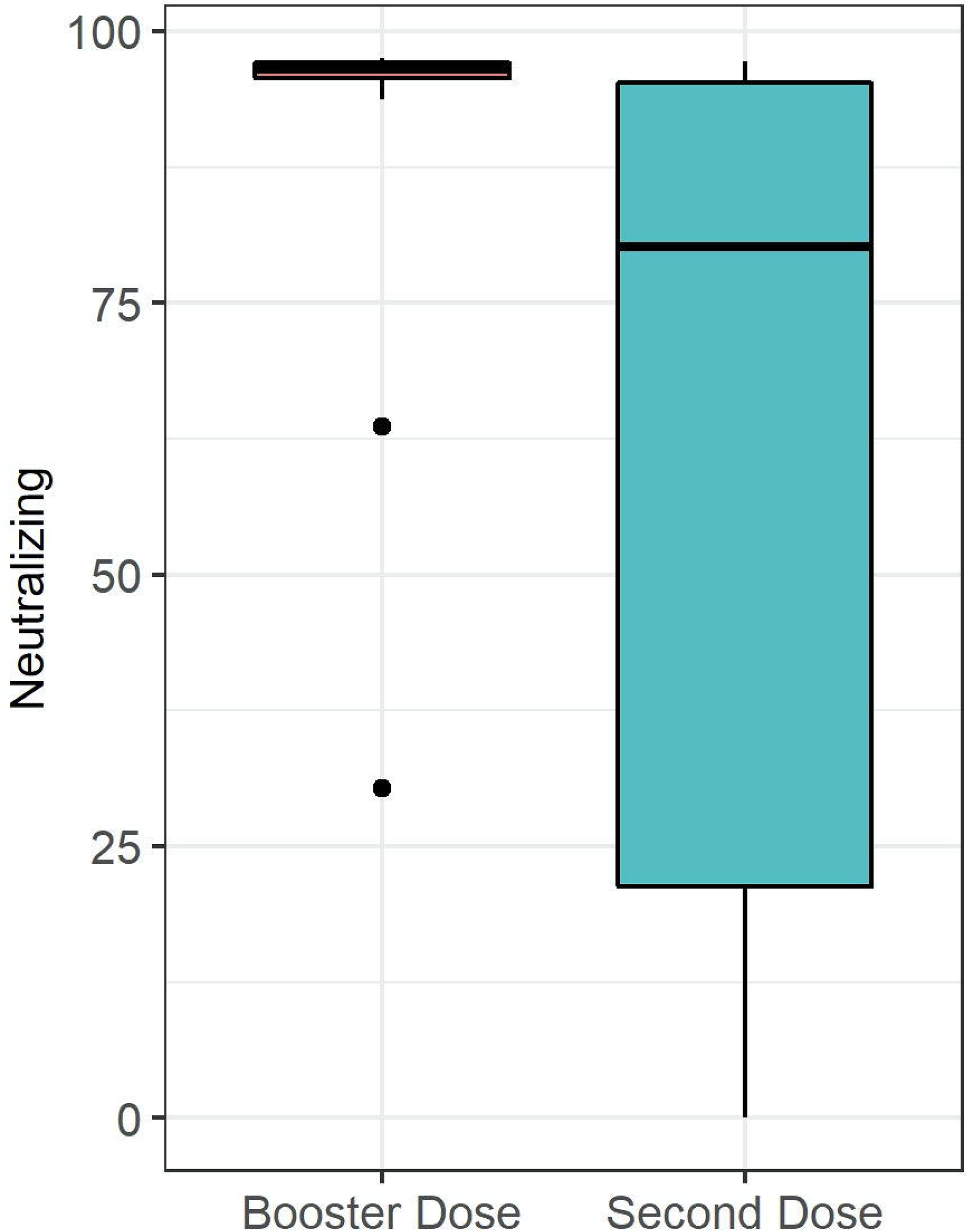
Box plot illustrating SARS-CoV-2-neutralizing antibody concentrations in the infliximab combination therapy after second and third dose.

The percentage of patients who achieved positive SARS-CoV-2 IgG levels in patients who received third (booster) dose was 96.3% (78 out of 81), whereas the percentage of patients with positive SARS-CoV-2 IgG levels (>31.5 BAU/mL) in patients who received second dose only was 86.4% (70 out 81) p = 0.026. The percentage of patients who received third (booster) dose and achieved positive SARS-CoV-2-neutralizing antibody level was 100%, whereas the percentage of patients was 88.9% (72 out 81) in the second dose group (p= 0.009). Finally, four patients had 0 neutralizing antibody level after the second dose. (see figure 4)

**Figure 4.**
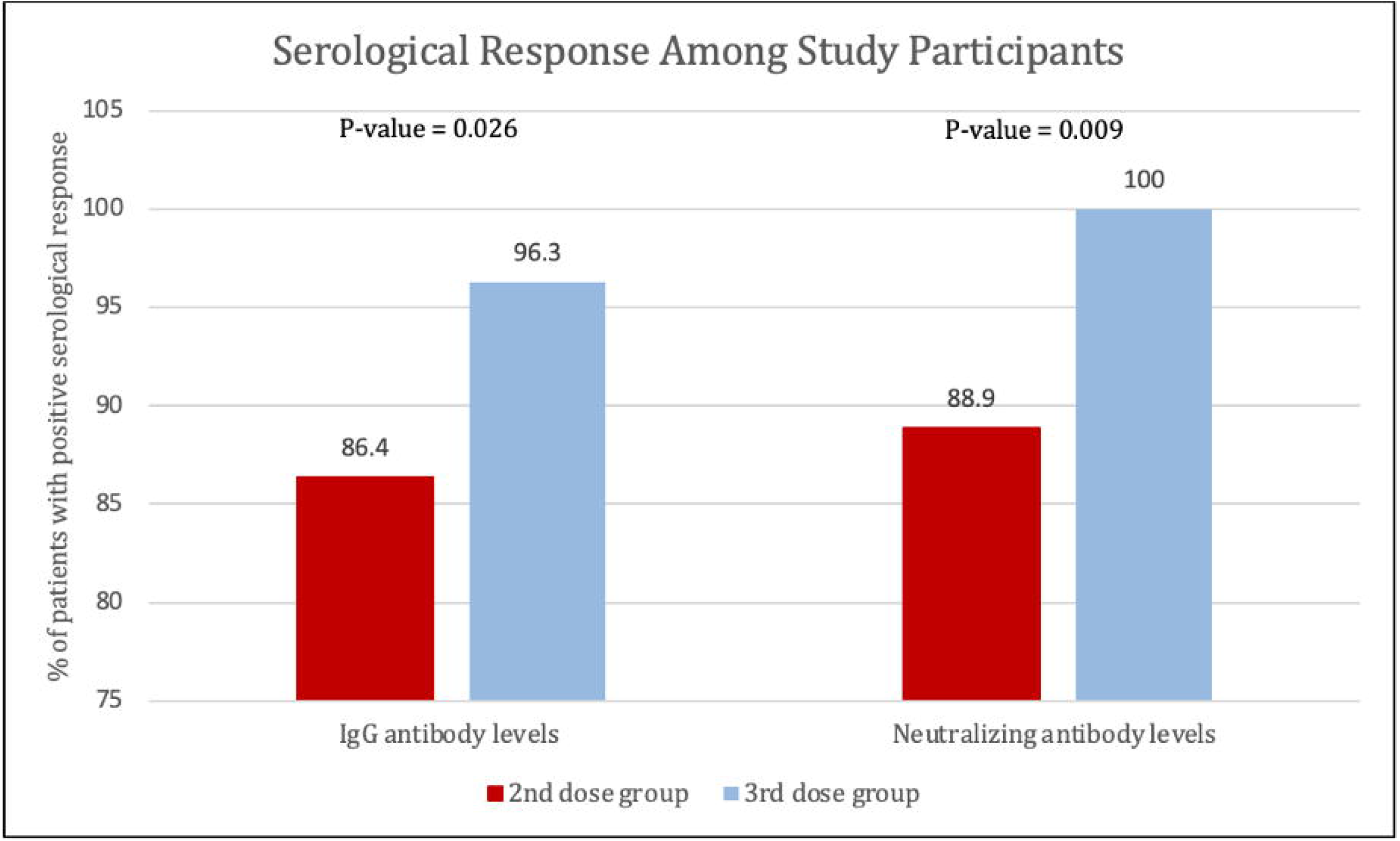
Percentage of patients with positive serological response to second and third dose of BNT162b2, defined by SARS-CoV-2 IgG antibody levels of >31.5 BAU/mL or SARS-CoV-2-neutralizing antibody levels >20%.

## Discussion

In this study we found that all patients with IBD on infliximab combination therapy had positive SARS-CoV-2 IgG and neutralizing antibody concentrations 40-45 weeks post BNT162b2 vaccination regardless of whether patients received a booster dose or not. However, SARS-CoV-2 IgG and neutralizing antibody concentrations were lower in patients who received 2 doses only compared to patients who received a third dose. Our study highlighted the importance of booster dose in the IBD population. Levin et al^17^ showed that six months after receipt of the second dose of the BNT162b2 vaccine, humoral response was substantially decreased, among persons 65 years of age or older, and among persons with immunosuppression. Levin et al also showed that SARS-CoV2 IgG and neutralizing antibody levels were lower in older men and participants with immunosuppression.

Another important finding of our study is that the percentage of participants who received third (booster) dose and achieved positive SARS-CoV-2-neutralizing antibody level was significantly higher at 100%, relative to the percentage of patients who received second dose only (88.9%). A study by Bergwerk et al recently reported a correlation between neutralizing antibody titers and infectivity.^18^ Similar to our study, Deepak et al^19^ found that 88.7% (118 of 133) of patients with chronic inflammatory disease such as IBD and rheumatoid arthritis treated with immunosuppressive medications had antibody responses after 2 doses of the BNT162b2 or mRNA-1273 vaccine; however, in many cases, antibody levels were lower than in immunocompetent participants. These studies suggested that although the specific threshold of antibody levels that can confer protection against breakthrough infection is still unclear, neutralizing antibodies may still be used as to determine efficacy and protection of vaccine.

The effect of tumor necrosis factor (TNF) antagonists on serological responses of COVID-19 vaccination is becoming well documented in the literature. One study^20^ recruited 362 patients with IBD treated with different immunosuppressive treatment regimens and 121 healthy controls. The authors found that patients being treated with infliximab or tofacitinib had lower anti-SARS-CoV-2 spike protein antibody concentrations after two doses of vaccine than did healthy controls. On the other hand, reductions in antibody responses were not observed in patients with IBD being treated with thiopurines, ustekinumab, or vedolizumab compared to control participants. Surprisingly, patients treated with infliximab 10-times reduction in anti-SARS-CoV-2 spike protein antibody concentrations relative to control group. Similarly, another study by Shehab et al recruited 116 patients with IBD receiving infliximab combination therapy. Authors concluded that in patients with IBD receiving infliximab combination therapy, SARS-CoV2 IgG, IgA and neutralizing antibody levels were lower compared with healthy participants.^14^ Finally, Melmed et al published a study that assessed antibody titers in adults with IBD who received mRNA SARS-CoV-2 vaccination.^21^ The study included 582 participants with IBD receiving different immunosuppressive therapies. Authors found that mean SARS-CoV-2 antibody levels at 8 weeks were the highest among those treated with anti-integrin and anti–interleukin-12/23, and lowest among those treated with anti-TNFs combination therapy or corticosteroids; however, the study was not powered to assess differences across medication subgroups. In our study we also noticed wide variability in range of IgG and Neutralizing Abs, which could be attributed to age and co-morbidity differences among patients. However, the ranged was narrow after the booster dose, which support recommendation for a third dose.

Our results build on growing literature confirming that most patients with IBD can mount humoral responses after second dose of SARS-CoV-2 vaccination, with a small proportion generating poor or no response, which provides justification for current recommendations for this population to receive a booster dose of BNT162b2 vaccine. In addition, our predefined inclusion and exclusion criteria lowers the risk of confounding bias, and patients were equally distributed in terms of demographics characteristics such as age, sex, and BMI.

This study has some limitations. We cannot be definitive that some of the included patients might have had silent COVID-19 at the time recruitment which could provoked higher serological response to vaacine,^22^ however; we did PCR testing before each vaccine dose and also excluded any patients with current or previous symptoms of COVID-19. In addition, we only assessed positive IgG and neutralizing antibody. However, cellular immunity may also play a role in vaccine efficacy. Finally, we investigated infliximab with azathioprine or 6-mercaptopurine only. Further studies are needed to investigate the effect of other immunomodulators.

## Conclusion

Most patients with IBD on infliximab combination therapy had positive SARS-CoV-2 IgG and neutralizing antibody concentrations 40-45 weeks post BNT162b2 vaccination. However, SARS-CoV-2 IgG and neutralizing antibody concentrations were lower in patients who received 2 doses only compared to patients who received a third dose. A Longer follow up study is needed to evaluate decay in antibodies overtime.

## Data Availability

All data produced in the present study are available upon reasonable request to the authors

## Declaration

All authors declare no conflict of interest

## Acknowledgment

**none**

## Contribution to the field statemen

BNT162b2 vaccine has been shown to be effective in decreasing COVID-19 associated morbidity and mortality. It has also been recommended for patients with inflammatory bowel disease (IBD). In this study, we aimed to evaluate if a booster (third) dose is needed for patients with IBD on infliximab combination therapy as these patients my potentially have reduced immune response to BNT162b2 vaccine compared to health individuals.

